# Intimate partner violence and unmet need for family planning among currently married Afghan women aged 18-49 years: findings from a nationally representative survey in Afghanistan

**DOI:** 10.1101/2021.10.20.21265274

**Authors:** Omid Dadras, Takeo Nakayama, Masahiro Kihara, Masako Ono-Kihara, Chamnong Thanapop, Phiman Thirarattanasunthon, Kasemsak Jandee

## Abstract

**Background:** Intimate Partner Violence (IPV) is a serious public health issue, particularly in poor-resourced countries. It has been linked to a range of adverse health outcomes. In this study, we explored the prevalence and the relationship between the IPV and unmet need for family planning and identified the associated sociodemographic factors among a nationally representative sample of married Afghan women aged 18-49 years in Afghanistan.

**Methods:** This study used the data from Afghanistan Demographic and Health Survey (DHS) conducted in 2015. The data for IPV, unmet need for family planning, and sociodemographic characteristics of Afghan women aged 18-24 were extracted from the available databases. Sampling weights and survey design were accounted for in bivariate and multivariate analyses using the STATA software version 14. The significant alpha level was determined at p-value < .05

**Results:** More than half of the study population, with a prevalence of 55.89%, experienced some types of intimate partner violence during the last 12 months. Unmet need for family planning was documented in less than a third of the population. Illiterate employed women from poorer families who were living in the rural areas were more likely to suffer from any type of IPV. Moreover, those from Pashtun (OR = 2.12, 95% CI:1.30-3.45), Tajik (OR = 1.69, 95% CI:1.06-2.71), and Pashai (OR = 2.23, 95% CI:1.17-4.26) ethnic groups had a higher chance of being a victim of any type of violence compared to the reference group (Turkmen). Illiterate women (OR = 1.37, 95% CI:1.02-1.83) with more than 5 pregnancy experiences (OR = 1.44, 95% CI: 1.19-1.74) had more unmet needs for family planning compared to the reference group. The unmet needs were almost 40% and 30% less likely to be observed among women from Pashtun and Tajik ethnic backgrounds compared to the reference group (Turkmen), respectively. The likelihood of having unmet needs was 30% less in those women who suffered from any type of violence.

**Conclusion:** IPV is an important predictor of several adverse health outcomes. The findings of present study portray the disastrous situation of Afghan women right violation and violence against them in a conflict setting in one the poor-resourced countries in the world and communicate an important message to the international communities and human right advocate to take immediate actions to mitigate the current situation and prevent the violence against Afghan women to improve the integrity of their reproductive health.

## Introduction

Intimate Partner Violence (IPV) is a serious public health issue, particularly in poor-resourced countries(1). It not only involves physical, emotional, and sexual violence but also the controlling behavior of the partner (2). Intimate partner violence (IPV) has been linked to a range of adverse health outcomes such as poor mental health (1, 3), adverse pregnancy and reproductive outcomes such as abortion, unintended pregnancy, and sexually transmitted diseases (1, 4-6). IPV is a common phenomenon in conflict and post-conflict settings and Afghanistan is one of the countries with a high prevalence of IPV in women (7, 8). Research indicates an estimated 50% of lifetime IPV among Afghan women (9). High gender inequality, a patriarchal society, ongoing internal conflict, early marriage, poverty, and low literacy account for the high prevalence of IPV toward women in most of the poor-resources countries (9-11).

A variety of social, cultural, and economic factors could influence women’s decisions and autonomy regarding family planning and contraceptive use. It has been shown that IPV could influence the use of contraceptives and lead to unmet need family planning among the victims; however, the results are contradictory. Unmet need for family planning is defined as the lack of use of any contraception method among fecund and sexually active women who report not wanting any more children or wanting to delay the next child (12). It is likely that women experiencing IPV use and seek less contraception out of fear of additional violence from their partners. They may also be more vulnerable due to inability in negotiating their reproductive need and right with their partners compared to other women. On the other hand, women who experienced IPV may be more willing to use contraception to avoid pregnancy and born a child into abusive marriages or relationships. In addition, to protect themselves from contracting sexually transmitted diseases and HIV/AIDs from a risky partner, they may also prefer and seek using contraceptives (2).

Several studies have explored the possible impact of IPV on contraceptive use among sufferers; however, the results are inconsistent. While, some studies from Colombia, Jordan, India, and Nigeria reported higher unmet needs for family planning among women who experienced IPV (13-17), other studies from Sub-Saharan Africa, Nigeria, and Kenya suggested higher use of contraceptives among women suffering from IPV (2, 4, 18). There are also some studies that reported no association between IPV and unmet need for family planning (19-21). Therefore, it is important to understand the relationship between IPV and the unmet need for family planning as well as the influencing sociodemographic factors that shape this relationship, especially in countries such as Afghanistan with one the highest rate of IPV and unmet need for family planning in the world.

Given this background, we aimed to examine the relationship between IPV and the unmet need for family planning among married Afghan women aged 18-49 years old in Afghanistan using the secondary data from the Demographic and Health survey 2015 (DHS 2015). We assume that facing the different challenges and opportunities, women could respond differently at each stage of their reproductive lives to the influencing factors on contraceptive use, including IPV.

## Methods

### Study design

This study used the data from Afghanistan Demographic and Health Survey (DHS) conducted in 2015 (ADHS 2015). It is a nationally representative survey implemented by the Central Statistics Organization (CSO) in collaboration with the Afghanistan Ministry of Public Health (MoPH) and funded by the United States Agency for International Development (USAID).

### Participants

The Afghanistan DHS 2015 employed a stratified two-stage sample design to estimate the key indicators at the national level, in urban and rural areas, and for each of the 34 provinces of Afghanistan. At the first stage, 950 clusters consisting of EAs (enumeration areas) were selected; 260 in urban areas and 690 in rural areas. An EA is a geographic area consisting of a convenient number of dwelling units that serve as counting units for the census provided by the Central Statistics Organization (CSO). At the second stage, 25,650 households were selected through an equal probability systematic selection process. Due to the approximately equal sample size in each province, to obtain a representative estimate at the national level weighting components were calculated and applied. All the women aged 15-49 years who were either permanent residents of the selected households or visitors who stayed in the households the night before the survey were recruited after informed consent. However, we assumed that the reasons behind the unmet family planning for married women aged 15-18 years could be different as the marriage in this age group is defined as child marriage; therefore, we excluded them from our analysis. A more detailed description of the sampling procedure is reported in the Afghanistan DHS 2015 final report (22). Although 29,033 eligible women aged 18-49 years old were interviewed in Aghanistan DHS 2015 (96.8% response rate), following the WHO guidelines “Putting Women First: Ethical and Safety Recommendations for Research on Domestic Violence against Women” World Health Organization, 2001, only one woman from all eligible women in the household was randomly selected for the interview. Additionally, due to the sensitive nature of the questions related to IPV, some of the selected women avoided the interview; thus, only 20,593 women were included in the final analysis.

### Scales and measurement

A standard DHS questionnaire including the questions related to the demographic and health issues was administered to the eligible women. The questionnaire collected data on women’s demographic characteristics, family planning, fertility preferences, child health, sexually transmitted diseases, marriage and sexual behavior, and domestic violence (23). However, for the current study, our primary interest was questions related to intimate partner violence and unmet needs for family planning.

Exposure to IPV was measured using a modified version of the Conflict Tactic Scale (CTS) (24), in which any experiences of IPV during the past 12 months were sought in detail. Physical violence (PV) was defined as being slapped, kicked, bitten, pushed, punched, choked, burnt on purpose, or assaulted using a knife or other weapons. Sexual violence (SV) was defined as forced sexual intercourse; degrading or humiliating sexual acts, or engaging in sexual intercourse out of fear. Emotional violence (EV) was described as exposure to verbal abuse or insults; made to feel bad about oneself; belittled in front of other people; scared or intimidated; threatened with violence or confronted with threats of harm. The responses were dichotomized into 1= “yes” and 0= “no”.

Unmet need was defined using the revised method and includes the unmet need for limiting (i.e. women whose most recent pregnancy was not wanted at all, fecund women who did not use contraception despite their desire to have no more children, women who were postpartum amenorrheic for 2 years following an unwanted birth and were not using contraception) and spacing (i.e. women whose most recent pregnancy was not wanted initially but wanted later, fecund women not using contraception who were undecided when/if they wanted a to have a child or who wanted a child 2+ years later, and women who were postpartum amenorrheic for 2 years following a mistimed birth and were not using contraception) (25). Many of these variables had dichotomous response alternatives (i.e., ‘yes’ or ‘no’ responses). Other explanatory variables included in the model were woman’s age, education, employment, parity, wealth quintile, place of residence, ethnicity, and husband’s age and education,

### Statistical analysis

Analysis was performed using STATA version 14. Due to the strict Islamic culture of the Afghan community and the prohibited extramarital sex, we assumed that seeking family planning services among unmarried Afghan women could be erroneously underestimated and yield biased estimates; therefore, only married women were included in our analysis. Descriptive statistics were employed to describe the participant’s characteristics and estimate the prevalence of unmet family planning and IPV among women of different age groups, accounting for the sampling weight and design to obtain nationally representative estimates at the national level.

Logistic regression analysis was used for two purposes: first, to estimate the odds of IPV and its component as well as the unmet needs for family planning across the different sociodemographic factors. Second, to generate unadjusted and adjusted odds ratios (ORs) and 95% confidence intervals (CIs) for the use of contraception across the different types of intimate partner violence among married Afghan women. In the adjusted model, women who did not report experiencing IPV were the referent group. Sampling weight and design were defined and accounted for using the STATA command “svy” in all analyses. All tests of hypothesis were 2-tailed, with a type 1 error rate fixed at 5%.

### Ethical consideration

DHS conforms to the US. Department of Health and Human Service regulations concerning human rights. In addition, this survey was approved by the Institutional Review Board (IRB) of the Afghanistan Ministry of Health (MoH). Besides, we sought the ethical necessities of this study from the Walailak Univerity Ethics Committee (WUEC). We also sought permission from the DHS website and filled a request to access and download the data.

## Results

A total number of 21361 married Afghan women aged 18-49 years were selected to be interviewed for assessing the intimate partner violence from whom 642 women could not complete the survey due to privacy reasons and 126 women for other reasons not specified; therefore, 20593 (96.4% response rate) women completed the survey and data were available for them.

### 1) The prevalence of IPV and unmet need for FP

More than half of the study population, with a prevalence of 55.89%, experienced some types of intimate partner violence during the last 12 months. Unmet need for family planning was documented in less than a third of the population. Almost a half suffered from physical assaults committed by husbands in the last 12 months. An estimated 7.54% of women appeared to experienced sexual violence at the same time.

### 2) Sociodemographic characteristics of married Afghan women

There was an almost equal number of Aghan women in age groups 18-24, 25-29, and ≥ 40; likewise for the age groups 30-34 and 35-39. The majority of participants were illiterate (85.23%), unemployed (87.75%), multipara (52.25%), and were living in rural areas (75.20%). Most of the women belonged to poor families (42.28%). More than two-thirds of participants were from Pashtun (39.21%) and Tajik (32.71%) ethnic backgrounds. Almost all husbands were employed but only less than half of them were literate (42.36%).

### 3) The sociodemographic determinants of IPV

#### 3.1) Physical violence determinants

Compared to the youngest age group (18-24), other age groups were experiencing more physical violence by their husband in the last 12 months (Table 3). In addition, illiterate women (OR = 2.02, 95% CI: 1.70, 2.39), those with higher parity, those from poor (OR = 1.41, 95% CI:1.18, 1.69) and middle-income (OR = 1.59, 95% CI:1.28, 1.96) families, those living in rural areas (OR = 1.75, 95% CI: 1.42, 2.14), and those from Pashtun ethnic background were more likely to suffer from physical violence during the last 12 months. Moreover, it appeared that illiterate husbands (OR = 1.54, 95% CI: 1.33, 1.79) from older age groups committed more physical assaults against their wives.

**Table 1.** The Sociodemographic Characteristics of married Afghan women aged 18-49 years, ADHS 2015

| Variables | N (%) |
| --- | --- |
| Age groups (years) |  |
| 18-24 | 4703 (22.84) |
| 25-29 | 4605 (22.36) |
| 30-34 | 3468 (16.84) |
| 35-39 | 3436 (16.69) |
| ≥ 40 | 4381 (21.27) |
| Education |  |
| Illiterate | 17551 (85.23) |
| Literate | 3042 (14.77) |
| Employment |  |
| Employed | 2522 (12.25) |
| Unemployed | 18071 (87.75) |
| Parity |  |
| 1 | 1893 (9.91) |
| 2-4 | 7233 (37.85) |
| >5 | 9985 (52.25) |
| Husband's age (year) |  |
| 18-24 | 1794 (8.75) |
| 25-34 | 7275 (35.49) |
| 35-44 | 6276 (30.61) |
| >45 | 5156 (25.15) |
| Husband Education |  |
| Illiterate | 11754 (57.64) |
| Literate | 8639 (42.36) |
| Wealth index |  |
| Poor | 8706 (42.28) |
| Middle | 4411 (21.42) |
| Rich | 7476 (36.30) |
| Place of Living |  |
| Urban | 5107 (24.80) |
| Rural | 15486 (75.20) |
| Ethnicity |  |
| Pashtun | 8062 (39.21) |
| Tajik | 6726 (32.71) |
| Hazara | 2149 (10.45) |
| Uzbek | 1568 (7.63) |
| Turkmen | 482 (2.34) |
| Nuristani | 632 (3.07) |
| Baloch | 268 (1.30) |
| Pashai | 343 (1.67) |
| Other | 330 (1.61) |
| Total | 20593 (100) |

**Table 2.**
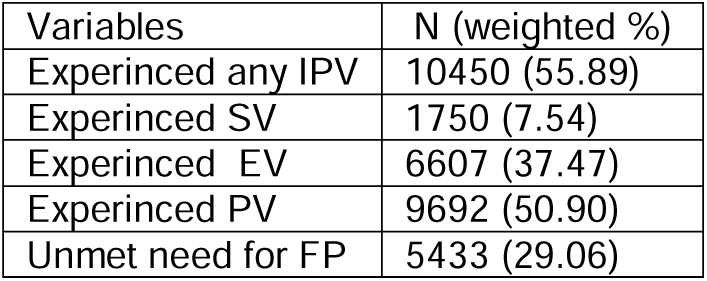
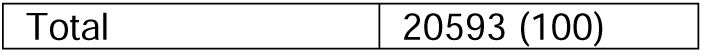
Prevalence of the different components of spousal violence among married Afghan women aged 18-49 years, ADHS 2015

| Variables | N (weighted %) |
| --- | --- |
| Experienced any IPV | 10450 (55.89) |
| Experienced SV | 1750 (7.54) |
| Experienced EV | 6607 (37.47) |
| Experienced PV | 9692 (50.90) |
| Unmet need for FP | 5433 (29.06) |
| Total | 20593 (100) |

**Table 3.** The odds of IPV and unmet need for FP across different sociodemographic factors of married Afghan women aged 18-49 years, ADHS 2015

| Variables | Experienced PA | Experienced SV | Experienced EV | Experienced any IPV | Unmet need for FP |
| --- | --- | --- | --- | --- | --- |
|  | OR (95% CI) | OR (95% CI) | OR (95% CI) | OR (95% CI) | OR (95% CI) |
| Age groups (years) |  |  |  |  |  |
| 18-24 | Ref | Ref | Ref | Ref | Ref |
| 25-29 | 1.49 (1.30-1.72) | 1.37 (1.08-1.75) | 1.51 (1.28-1.79) | 1.52 (1.33-1.73) | 1.23 (1.06-1.43) |
| 30-34 | 1.56 (1.34-1.80) | 1.24 (.96-1.56) | 1.41 (1.20-1.67) | 1.41 (1.23-1.63) | 1.37 (1.15-1.64) |
| 35-39 | 1.68 (1.48-1.90) | .99 (.76-1.29) | 1.45 (1.23-1.71) | 1.51 (1.31-1.76) | 1.32 (1.04-1.68) |
| ≥ 40 | 1.87 (1.62-2.16) | 1.08 (.86-1.26) | 1.68 (1.41-2.09) | 1.73 (1.51-1.97) | 1.18 (.96-1.46) |
| Education |  |  |  |  |  |
| Illiterate | 2.02 (1.70-2.39) | 1.77 (1.26-2.49) | 1.57 (1.36-1.82) | 1.73 (1.45-2.06) | 1.37 (1.02-1.83) |
| Literate | Ref | Ref | Ref | Ref | Ref |
| Employment |  |  |  |  |  |
| Employed | 1.20 (.96-1.51) | 1.74 (1.32-2.30) | 1.27 (0.99-1.62) | 1.34 (1.05-1.71) | .87 (.67-1.12) |
| Unemployed | Ref | Ref | Ref | Ref | Ref |
| Parity |  |  |  |  |  |
| 1 | Ref | Ref | Ref | Ref | Ref |
| 2-4 | 1.34 (1.13-1.59) | 1.08 (.81-1.45) | 1.28 (1.09-1.52) | 1.27 (1.08-1.49) | 1.16 (.96-1.42) |
| >5 | 1.60 (1.34-1.92) | 1.13 (.84-1.52) | 1.39 (1.19-1.62) | 1.42 (1.21-1.65) | 1.44 (1.19-1.74) |
| Husband's age (year) |  |  |  |  |  |
| 18-24 | Ref | Ref | Ref | Ref | Ref |
| 25-34 | 1.56 (1.28-1.90) | 1.16 (.81-1.57) | 1.50 (1.25-1.79) | 1.63 (1.36-1.96) | 1.19 (.99-1.43) |
| 35-44 | 1.71 (1.45-2.02) | 1.07 (.77-1.48) | 1.72 (1.44-2.05) | 1.72 (1.44-2.07) | 1.22 (1.01-1.47) |
| >45 | 2.01 (1.66-2.43) | .93 (.67-1.29) | 1.91 (1.60-2.29) | 1.95 (1.63-2.32) | 1.28 (.99-1.64) |
| Husband Education |  |  |  |  |  |
| Illiterate | 1.54 (1.33-1.79) | 1.13 (.92-1.38) | 1.33 (1.17-1.52) | 1.45 (1.24-1.69) | 1.04 (.92-1.18) |
| Literate | Ref | Ref | Ref | Ref | Ref |
| Wealth index |  |  |  |  |  |
| Poor | 1.41 (1.18-1.69) | 1.61 (1.17-1.23) | 1.06 (.89-1.27) | 1.19 (.99-1.43) | 1.32 (1.09-1.60) |
| Middle | 1.59 (1.28-1.96) | .90 (.63-1.28) | 1.17 (1.01-1.37) | 1.36 (1.08-1.71) | 1.22 (.96-1.55) |
| Rich | Ref | Ref | Ref | Ref | Ref |
| Place of Living |  |  |  |  |  |
| Urban | Ref | Ref | Ref | Ref | Ref |
| Rural | 1.75 (1.42-2.14) | 1.19 (.77-1.82) | 1.09 (.93-1.29) | 1.38 (1.13-1.69) | 1.06 (.89-1.27) |
| Ethnicity |  |  |  |  |  |
| Pashtun | 1.75 (1.08-2.82) | 1.95 (0.86-4.38) | 4.10 (2.69-6.25) | 2.12 (1.30-3.45) | .61 (.44-.83) |
| Tajik | 1.44 (.89-2.32) | 2.72 (1.21-6.09) | 3.86 (2.54-5.88) | 1.69 (1.06-2.71) | .70 (.51-.97) |
| Hazara | 1.29 (.76-2.17) | 3.25 (1.30-8.13) | 3.17 (2.02-4.98) | 1.55 (.93-2.58) | .83 (.57-1.21) |
| Uzbek | 1.31 (.77-2.24) | .54 (.19-1.58) | 2.62 (1.57-4.38) | 1.66 (.97-2.84) | .88 (.60-1.31) |
| Turkmen | Ref | Ref | Ref | Ref | Ref |
| Nuristani | 1.41 (.84-2.38) | 3.71 (1.61-8.58) | 1.77 (1.09-2.87) | 1.60 (.94-2.73) | .66 (.44-1.01) |
| Baloch | .64 (.20-1.99) | .30 (.05-1.75) | 1.52(0.59-3.89) | .67 (.21-2.09) | .95 (.34-2.64) |
| Pashai | 1.94 (1.04-3.60) | 1.94 (.68-5.55) | 3.91 (2.11-7.26) | 2.23 (1.17-4.26) | .94 (.61-1.43) |
| Other | 1.48 (.74-2.96) | 4.01 (1.55-10.35) | 2.19 (1.21-3.95) | 1.66 (.85-3.26) | 1.09 (.71-1.68) |

#### 3.2) Sexual violence determinants

The age group 25-29 was more likely to suffer from sexual violence compared to other age groups (Table 3). Moreover, illiterate (OR = 1.77, 95% CI:1.26, 2.49), employed (OR = 1.74, 95% CI:1.32, 2.30) women from poor families (OR = 1.61, 95% CI:1.17, 1.23) were more likely to experience sexual violence in the last 12 months. The odds of experiencing sexual violence were 2.72, 3.25, and 3.71 times higher among Tajik, Hazara, and Nuristani ethnicities compared to the reference group (Turkmen).

#### 3.3) Emotional violence determinants

All the age groups were more likely to experience emotional violence compared to the youngest age group (18-24). It appeared that illiterate women (OR = 1.57, 95% CI: 1.36, 1.82) with multiple pregnancy experiences (more than 2) from middle-income families experienced more emotional violence compared to the reference group. Furthermore, women from all ethnic backgrounds, except for the Baloch, were at a higher risk of being emotionally abused in the last 12 months by their husbands (Table 3).

#### 3.4) The determinants of intimate partner violence (any type)

The odds of experiencing any type of intimate partner violence was almost 1.5 times higher in all age groups compared to the youngest age group. Illiterate employed women from poorer families who were living in the rural areas were more likely to suffer from any type of IPV (Table 3). Moreover, those from Pashtun (OR = 2.12, 95% CI:1.30-3.45), Tajik (OR = 1.69, 95% CI:1.06-2.71), and Pashai (OR = 2.23, 95% CI:1.17-4.26) ethnic groups had a higher chance of being a victim of any type of violence compared to the reference group (Turkmen). In addition, illiterate husbands (OR = 1.45, 95% CI: 1.24-1.69) older than 25 years old were more likely to commit any type of violence against their wives (Table 3).

### 4) The sociodemographic determinants of unmet need for family planning

It appeared that women aged 25-39 are more likely to suffer from the unmet need for family planning. In addition, illiterate women (OR = 1.37, 95% CI:1.02-1.83) with more than 5 pregnancy experiences (OR = 1.44, 95% CI: 1.19-1.74) had more unmet needs for family planning compared to the reference group. The unmet needs were almost 40% and 30% less likely to be observed among women from Pashtun and Tajik ethnic backgrounds compared to the reference group (Turkmen), respectively. Moreover, older husband age was associated with the higher unmet need for family planning (Table3)

### 4) Intimate partner violence and unmet need for family planning

Table 4 shows both unadjusted and adjusted odds ratios and 95% confidence intervals for the relationship between different components of intimate partner violence and unmet need for family planning among Afghan women aged 18-49 years before and after adjustment for sociodemographic factors. As it is shown, those who suffered from physical and emotional violence were almost 30% less likely to have unmet needs for family planning compared to the reference group after adjustment for sociodemographic factors. Likewise, the likelihood of having unmet needs was 30% less in those women who suffered from any type of violence compared to those who did not.

**Table 4.**
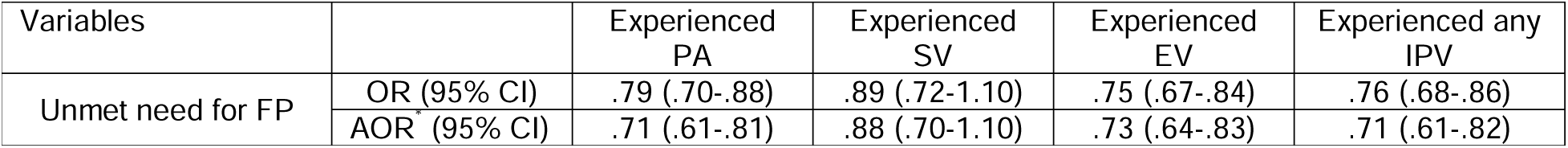
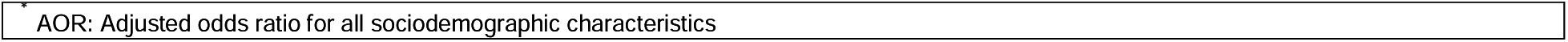
The association between different components of IPV with unmet need for family planning among married Afghan women aged 18-49 years, ADHS 2015

| Variables |  | Experienced PA | Experienced SV | Experienced EV | Experienced any IPV |
| --- | --- | --- | --- | --- | --- |
| Unmet need for FP | OR (95% CI) | .79 (.70-.88) | .89 (.72-1.10) | .75 (.67-.84) | .76 (.68-.86) |
|  | AOR (95% CI) | .71 (.61-.81) | .88 (.70-1.10) | .73 (.64-.83) | .71 (.61-.82) |

## Discussion

This is the first study that provides nationally representative estimates of intimate partner violence, unmet need for family planning, and associated sociodemographic factors among married Afghan women aged 18-49 years old living in Afghanistan. The study also determined the association between IPV and the unmet need for family planning. The results indicated that more than half (55.89%) of the married Afghan women aged 18-49 experienced some types of violence during the last 12 months which showed a decreasing trend from 73% in 2008 (8). It appeared that the woman in age groups older than 25 years are more likely to experience spousal violence compared to younger age groups. Likewise, illiterate husbands of older ages seem to commit more violent acts against their wives. The likelihood of spousal violence was often more among poor illiterate rural women with more pregnancy experiences. Sexual violence was more commonly observed among employed women. Women from Pashtun, Tajik, and Pashai ethnic groups were more likely to suffer from any type of spousal violence. The prevalence of unmet need for family planning was almost 30%. The middle age groups (25-39) were more likely to have unmet needs for family planning. Illiteracy and poverty were associated with higher unmet needs. It appeared that married women who experienced any type of spousal violence during the last 12 months had fewer unmet needs for family planning.

### IPV and unmet need for family planning

Intimate partner violence (IPV) is a common phenomenon in both conflict and post-conflict settings, but the possible health outcomes of IPV in such settings are understudied (26). It has been more than four decades that Afghan people are suffering from high levels of internal conflict and the country has one the highest prevalence of IPV among low-income countries (27). However, the possible outcomes of such violence on access to the contraceptive and unmet need for family planning are still under debate. Some studies suggested that the women’s less autonomy in decision making in an abusive relationship reduce the women’s access to contraceptive and family planning (17, 28, 29); while, there is some contradictory evidence that shows that it could increase the utilization of contraceptive and lead to less unmet need for family planning as the victimized women are more likely to seek contraceptive in order to prevent bringing a child into this abusive and hostile environment and protect herself against the sexually transmitted diseases that may be acquired from a risky partner (2, 4, 18). Similarly, in our study, we observed lower rates of unmet need for family planning among those who suffered from any type of IPV (physical and emotional) except for sexual violence. This may indicate that Afghan women in an abusive relationship are seeking more contraceptives to avoid conception with a child that may be harmed by the insecure and hostile environment. Therefore, policymakers in Afghanistan need to consider measures to mitigate these gender-based issues, particularly IPV, to improve Afghan women’s reproductive health and ensure their access to necessary family planning packages.

There are several factors that predict the incidence of spousal violence such as literacy, socioeconomic status, consanguinity, social norms, and culture (30, 31); however, they could be context-specific. For example, in the Muslim communities, the patriarchal structure of the families and man-dominant decision-making reduce the autonomy of the women. This not only influences women’s decision-making in seeking critical reproductive needs but also increases the chances of being physically and emotionally assaulted by a violent partner/husband at the time of disagreement (32). Furthermore, the women’s low literacy, poverty, and unemployment could further reduce their undependability and autonomy in life decision-making and increase the risk of spousal violence (31). Likewise, our study was conducted in a poor-resources strict Islamic setting in which the majority of women are illiterate and living in poverty and their human rights are often violated. Besides, evidence show that rigid traditional views and gender norms in Afghan society turned wife-beating into an acceptable act when women violate the gender norms and resist male wishes (33). This could also lead to under-reporting of IPV and suppress the help-seeking behaviors of victims among women in Afghanistan as the women might adhere to these rigid traditional gender roles to protect themselves from further abuse and spousal aggression (34). Furthermore, the sensitive nature of gender-based violence prevents Afghan women to disclose and share their experiences of such violence (33). Therefore, the international parties and human rights advocates should consider addressing these issues by formulating culturally sensitive interventions and policies to enhance the safety net for Afghan women and improve their help-seeking behavior in a safe and secure environment.

Besides the several adverse health outcomes of IPV, the unmet need for family planning which often leads to unwanted pregnancy, abortion, and STDs and harms the reproductive health of the victim is an important public health issue (17). Although we have found lower rates of unmet need for family planning among those suffering from IPV, the observed distribution of sociodemographic factors of the study population, particularly the low literacy and poverty, and their association with unmet needs are concerning and this once again highlighted the importance of these sociodemographic factors on adverse health outcomes especially among the rural population in poor-resourced countries. We found that not only the women’s but also the husband’s literacy is an important determinant of both IPV and the unmet need for family planning which was in line with previous studies (30, 35). The results also indicated higher rates of IPV among the Pashtun and Tajik ethnic groups, similar to the previous study in Afghanistan (9); however, to the best of our knowledge, we are the first study that documented the lower unmet need of family planning among married women aged 18-24 that belong to these ethnic groups. It is important as the majority of the Afghan population are either Pashtun or Tajik and a high rate of IPV could harm the mental and physical health of the women from these ethnic groups and should be considered and addressed in future policies and interventions.

## Limitations

Although this was the first study that provided nationally representative estimates for different components of IPV and the unmet need for family planning among married Afghan women aged 18-49 years in Afghanistan, some shortcomings need to be considered in interpreting the results of the present study. First, the sensitive nature of gender-based issues such as IPV and the fear of disclosure in the patriarchic Afghan society may lead to underreporting and underestimation of the IPV in this study. In addition, the socially desirable response is an inherent bias in such surveys and may distort the estimates of IPV an unmet need for family planning. Second, despite the high rates of physical and emotional violence, the rate of sexual violence seemed to be considerably low, this has been attributed to the misunderstanding of forced sex as a right and not a violent act for the husband by Afghan women and thus underreporting of sexual violence (34). Third, food insecurity has been documented as an important predictor of IPV (9); however, DHS 2015 did not collect the data on this variable and we could not include this variable in the analysis. Nonetheless, it has been shown the food insecurity is strongly associated with the wealth index of the household which is also a predictor of contraceptives use among women (36). Therefore, we are confident to account for the effect of food insecurity by adjusting for the effect of the wealth index in multivariate analysis.

## Conclusion

IPV is an important predictor of several adverse health outcomes. In this study, IPV was observed in more than half of the married Afghan women aged 18-49 years and almost 30% reported unmet needs for family planning. We have found an inverse relationship between IPV and the unmet need for family planning. This could be due to the reluctance of the victim woman to bring a child to the hostile environment and protect herself from acquiring the sexually transmitted diseases that could be transmitted from a risky partner. Low literacy and poverty were some of the main determinants of the IPV and unmet need for family planning. We also observed higher rates of IPV among Pashtun and Tajik ethnic groups; conversely, the rate of unmet need for family planning was low among them. The findings portray the disastrous situation of Afghan women’s right violation and violence against them in a conflict setting in one the poor-resourced countries in the world and communicate an important message to the international communities and human right advocate to take immediate actions to mitigate the current situation and prevent the violence against Afghan women to improve their reproductive health.

## Data Availability

The DHS questionnaire that collected the data in Afghanistan's demographic and health survey in 2015 could be reached at DHS official website (https://www.dhsprogram.com). The dataset that was used in this study could be available upon a reasonable request and with the permission of the Walailak University ethical board.

## Authors contribution

OD wrote the research protocol and contributed to the data analysis and writing the manuscript. TN, MK, MOK provided critical feedback and comments on the data analysis and results and help in preparing the final draft. KJ, PT, and CT contributed to the writing of the research protocol, obtaining ethical approval, preparing the final draft.

## Acknowledgment

We would like to thank Prof. SeyedAhmad SeyedAlinaghi at the Iranian Center for HIV/AIDS Research, Tehran University of Medical Science, Iran, and Dr. Fateme Dadras at the Department of Gynecology and Obstetrics, Tehran University of Medical Science, Iran, for their feedback and advice on the methods and results.

